# Detection of Alteration in Carotid Artery Volumetry Using Standard-of-care Computed Tomography Surveillance Scans Following Unilateral Radiation Therapy for Early-stage Tonsillar Squamous Cell Carcinoma Survivors: A Cross-Sectional Internally-Matched Carotid Isodose Analysis

**DOI:** 10.1101/2024.02.03.24302288

**Authors:** Efstratios Koutroumpakis, Mohamed A. Naser, Abdallah Sherif Radwan Mohamed, Salman A. Eraj, Andrea Jarre, Jay C. Shiao, Mona Kamal, Subha Perni, Jack P. Phan, William H. Morrison, Steven J. Frank, G. Brandon Gunn, Adam S. Garden, Anita Deswal, Jun-ichi Abe, David I. Rosenthal, Elie Mouhayar, Clifton D. Fuller

## Abstract

**Purpose:** Radiation induced carotid artery disease (RICAD) is a major cause of morbidity and mortality among survivors of oropharyngeal cancer. This study leveraged standard-of-care CT scans to detect volumetric changes in the carotid arteries of patients receiving unilateral radiotherapy (RT) for early tonsillar cancer, and to determine dose-response relationship between RT and carotid volume changes, which could serve as an early imaging marker of RICAD.

**Methods and Materials:** Disease-free cancer survivors (>3 months since therapy and age >18 years) treated with intensity modulated RT for early (T1-2, N0-2b) tonsillar cancer with pre- and post-therapy contrast-enhanced CT scans available were included. Patients treated with definitive surgery, bilateral RT, or additional RT before the post-RT CT scan were excluded. Pre- and post-treatment CTs were registered to the planning CT and dose grid. Isodose lines from treatment plans were projected onto both scans, facilitating the delineation of carotid artery subvolumes in 5 Gy increments (i.e. received 50-55 Gy, 55-60 Gy, etc.). The percent-change in sub-volumes across each dose range was statistically examined using the Wilcoxon rank-sum test.

**Results:** Among 46 patients analyzed, 72% received RT alone, 24% induction chemotherapy followed by RT, and 4% concurrent chemoradiation. The median interval from RT completion to the latest, post-RT CT scan was 43 months (IQR 32-57). A decrease in the volume of the irradiated carotid artery was observed in 78% of patients, while there was a statistically significant difference in mean %-change (±SD) between the total irradiated and spared carotid volumes (7.0±9.0 vs. +3.5±7.2, respectively, p<.0001). However, no significant dose-response trend was observed in the carotid artery volume change withing 5 Gy ranges (mean %-changes (±SD) for the 50-55, 55-60, 60-65, and 65-70+ Gy ranges [irradiated minus spared]: −13.1±14.7, −9.8±14.9, −6.9±16.2, −11.7±11.1, respectively). Notably, two patients (4%) had a cerebrovascular accident (CVA), both occurring in patients with a greater decrease in carotid artery volume in the irradiated vs the spared side.

**Conclusions:** Our data show that standard-of-care oncologic surveillance CT scans can effectively detect reductions in carotid volume following RT for oropharyngeal cancer. Changes were equivalent between studied dose ranges, denoting no further dose-response effect beyond 50 Gy. The clinical utility of carotid volume changes for risk stratification and CVA prediction warrants further evaluation.

## Introduction

Oropharyngeal cancer (OPC), including tonsillar cancer, is the most common form of head and neck cancer, accounting for over 54,000 new cases in the US in 2023 alone.(1) The rising prevalence of HPV infection is leading to an increase in OPC in younger patients (age <55), who have greater survival rates, with the majority living on for decades.(1) This increase in survival brings a greater focus to treatment-related side effects, with carotid artery stenosis (CAS) and resultant cerebrovascular accident (CVA), being among the most serious ones. The relative risk of transient ischemic attack (TIA) or CVA is at least doubled by head and neck radiotherapy (RT), the backbone of OPC treatment.(2) Despite the relatively high incidence of TIA and CVA among patients treated with neck RT (approximately 5% in 5 years and 10% in 10 years), evidence-based guidelines for the management of radiation-induced carotid artery disease (RICAD) are largely anecdotal.(3–6) Currently, a major obstacle to the development of such guidelines is the lack of a quantified dose-risk relationship. Historically, carotid arteries received a uniform RT dose; however, since the early 2000’s, RT techniques, such as intensity-modulated RT (IMRT) have delivered a variable, non-homogenous dose to the carotid arteries, making the already crude estimators of CAS so anachronistic as to be functionally non-usable for risk stratification. Furthermore, several imaging modalities such as carotid ultrasound, computed-tomography (CT) scans and magnetic resonance imaging have been utilized to detect structural changes that predict subsequent development of clinical CAS. However, there is no evidence-based guidance as to which patients will benefit by which imaging, resulting in a heterogeneous use of these modalities in clinical practice. Development of personalized surveillance programs for the detection of RICAD based on baseline clinical and imaging characteristics as well as RT dose characteristics remains an unmet need. Standard-of-care CT scans with contrast, optimized for soft tissue visualization for staging, are performed around the time of diagnosis of OPC as well as within 6 months of treatment conclusion for determination of response.(1) Furthermore, longer term follow up CT scans of the head and neck area are frequently performed in OPC survivors to detect cancer recurrence or therapy-related complications. While the soft-tissue visualization optimized CT scans with contrast generally available for tonsillar cancer patients are not ideal for study of the carotid arteries, sufficient resolution is still present to accurately define the vessel’s diameter from the CCA’s origin at the aorta to the ICA’s entry of the skull as compared to ultrasound (US). Information of these widely-available standard-of-care CT scans can be used to identify patients at risk of CAS and guide dedicated carotid artery imaging for the vulnerable patients.

In previous work, Carpenter et al. performed an assessment of US-detected carotid stenosis (defined as ≥50% luminal reduction on imaging) in 366 patients with prior RT. Using these criteria, no formal dose-response correlation could be defined.(7) However, US is limited by the lack of volumetric imaging across the entire carotid volume. Consequently, in a larger longitudinal series, Carpenter et al. recently demonstrated that nearly all dose-levels between 10-70 Gy were associated with CAS risk increase, echoing prior work by van Aken, which showed that in unscreened patients a bilateral volume receiving ≥10Gy (V10) was associated with ischemic stroke risk.(8, 9)

While substantive vascular disease (asymptomatic carotid stenosis >50%) and consequential (ischemic) late events have been demonstrated to have an association with low dose, we hypothesized, based on related prospective US assessment of unilateral carotids, that early (<5-year) volumetric changes could be potentially detectable using volumetric imaging, without referral for dedicated CT-or MRI-angiography. The aim of this study was to utilize standard-of-care oncologic surveillance CT scans of the head and neck area performed prior and after unilateral RT for early-stage tonsillar cancer and assess volumetric changes in the carotid arteries in association with RT dose levels encountered in clinical practice. We hypothesized that volumetric changes in the carotid arteries, before and after unilateral RT for OPC would differ between the irradiated and untreated side in an identifiable dose-response relationship, serving as internally-matched self-controls. Identifying vulnerable patients for CAS, utilizing standard-of-care surveillance CT scans and defining radiation-dose-response parameters associated with carotid injury would enable development of improved treatment guidelines and actionable dose-modification strategies that will lead to decreased radiation-associated TIA/CVAs, thereby reducing therapy-attributable morbidity and mortality and improving patient quality of life. The specific aims of the current study included: 1) Demonstration of the technical feasibility of standard-of-care CT head and neck scans to detect alteration in post-RT carotid volumes. 2) Evaluation of dose-response of local carotid injury (e.g. at the location of dose) through isodose mapping. 3) Generation of preliminary data for future prospective observational and interventional approaches to early carotid radioinjury.

## Methods

### Patient population and data collection

Disease-free cancer survivors (18 years or older at diagnosis) previously treated for oropharyngeal squamous cell carcinoma at the University of Texas M.D. Anderson Cancer Center from October 2016 to May 2017 were identified. We utilized an extant dataset of CT images and clinical measures collected under an IRB-approved retrospective chart review protocol. Patients who were treated with unilateral intensity modulated RT (IMRT for early (T1-2, N0-2b) tonsillar cancer were included. We elected to study patients with early tonsillar cancer since they have low-probability of cancer-specific mortality and because early tonsillar cancer is often amenable to unilateral RT. With both a treated and untreated artery, the untreated carotid artery serves as a control to account for carotid artery volume changes related to patient-specific atherosclerotic processes. By leveraging intra-patient matched carotid comparisons, the variability associated with inter-patient atherosclerotic, cardiovascular, genetic or environmental confounders is modulated more effectively than with case-control or propensity matched cohorts. Patients treated with definitive surgery, bilateral RT, or additional RT prior to post-RT CT scan were excluded. Patient demographic, tumor and treatment characteristics were collected.(10)

### Computed tomography scans

Pre-RT CT scans included contrast-enhanced CT scans of the neck prior to initiation of RT. Post-RT scans included the last available (most recent) contrast-enhanced CT scans of neck. Two CT scans (pre- and post-RT) were analyzed for each patient. Contrast-enhanced CT scans were exported from the institution PACS to VelocityAI (version 3.0.1) in DICOM format. The RT plan must have been available and was exported from Pinnacle^3^ (version 14.0) to VelocityAI. The ICA/CCA (internal carotid artery/common carotid artery) were manually segmented in VelocityAI from the base of the skull to its origin at the aorta as depicted in Figure 1. We employed image registration in VelocityAI to align the planning CT with the pre- and post-therapy contrast-enhanced CT images. This alignment facilitated the transfer of isodose lines from the treatment plan to the pre- and post-therapy CT images. Consequently, we could delineate subvolumes of the carotid artery that received incremental dose ranges of 5 Gy, varying from 50 to 70+ Gy, as illustrated in Figure 1.

**Figure 1:**
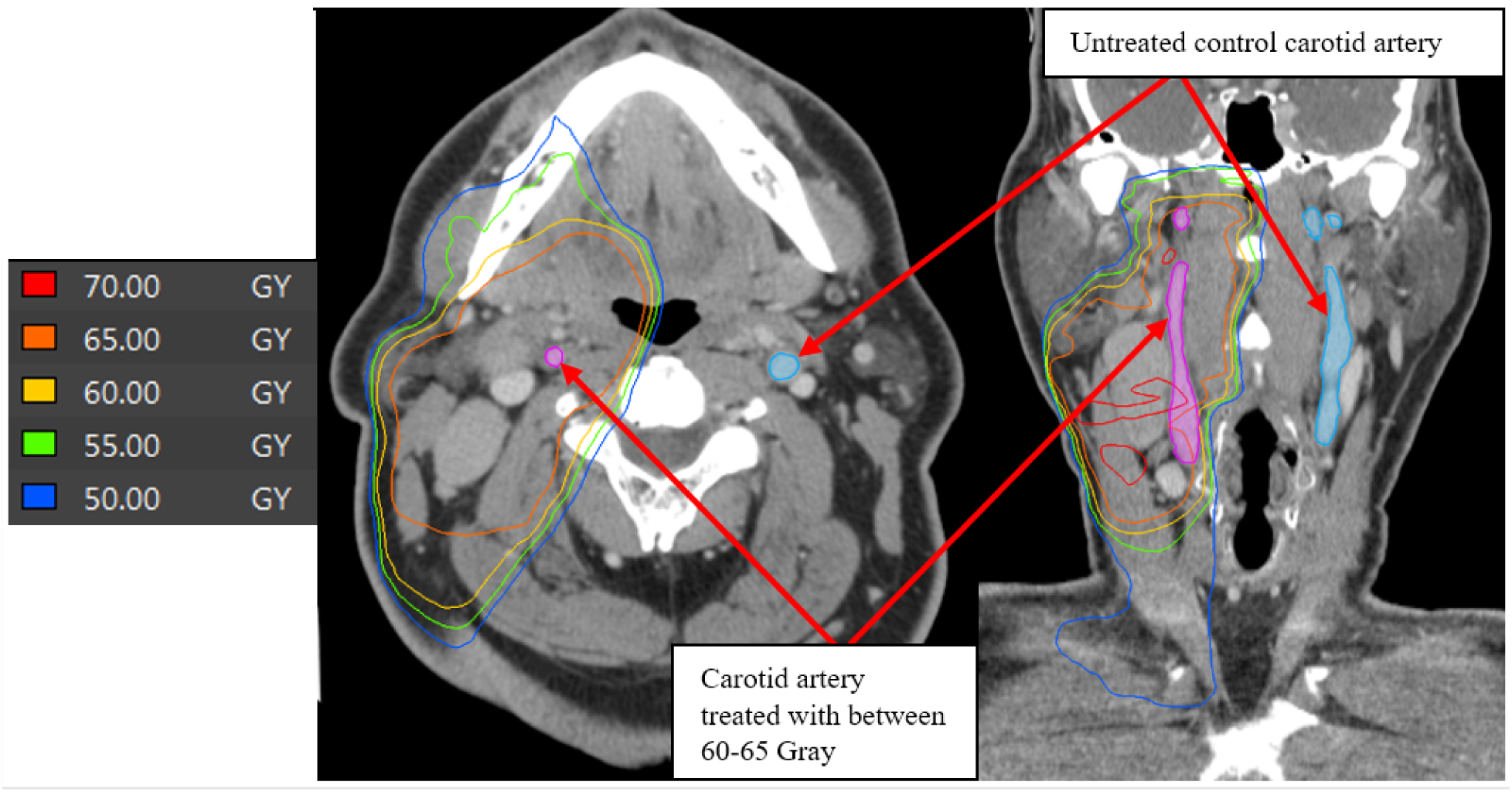
Axial and coronal views of carotid artery segmentation with isodose lines for each studied range.

### Statistical analysis

Descriptive statistics were used to summarize the patient, treatment, and tumor characteristics. The percent change in volume for each 5-Gy subvolume was calculated and compared to the contralateral side by the Wilcoxon rank sum test. Analysis of variance (ANOVA) was conducted in regards to the 5 Gy ranges and percent volume changes as continuous variables, in addition to literature-reported correlates of CAS (e.g. systemic therapy, neck dissection status, smoking status). Analysis was also conducted to determine if percent volume changes are related to CVA incidence.

## Results

A total of 46 patients were included in this analysis. Mean age ± SD was 54.5 ± 9.1 years, while 30% of patients were women and 96% white. Forty-four % of patients were either active (13%) or former (30%) smokers. None of the patients had CAS (>50% stenosis) at baseline. Most patients who were tested had human papilloma-associated (HPV+) disease (25/27; 93%). In terms of cancer therapy, 72% received RT alone, 24% received induction chemotherapy followed by RT, and 4% received concurrent chemoradiation. Neck dissection was performed in 20% of patients. Most patients received 66-69 Gy of RT dose (87%) divided in 30 or more fractions. Table 1 includes additional demographic and clinical characteristics.

**Table 1:** Demographic and basic clinical characteristics of 47 patients with early-stage tonsillar squamous cell carcinoma treated with unilateral neck radiation therapy.

|  |  |
| --- | --- |
|  | Total N=46 |
| Age, mean in years (SD) | 54.5 (9.1) |
| Female sex, N (%) | 14 (30.4) |
| White race, N (%) | 44 (95.6) |
| Hispanic ethnicity, N (%) | 2 (4.3) |
| Smoking history, N (%) | 20 (43.5) |
| - Active | 6 (13.0) |
| - Former | 14 (30.4) |
| HPV infection, N (%) | 25/27 (92.5) |
| Cancer Stage, N (%) |  |
| - T1N0 | 3 (6.5) |
| - T1N1-2b | 29 (63.0) |
| - T2N0 | 3 (6.5) |
| - T2N1-2b | 11(23.9) |
| Cancer therapy, N (%) |  |
| - RT alone | 33 (71.7) |
| - Induction chemotherapy + RT | 11 (23.9) |
| - Concurrent chemotherapy + RT | 2 (4.3) |
| - Neck dissection | 9 (19.6) |
| RT Fractions, N (%) |  |
| - 30 | 40 (87.0) |
| - 33 | 3 (6.5) |
| - 35 | 1 (2.2) |
| - 40 | 2 (4.3) |
| RT dose delivered, N (%) |  |
| - 60-64 | 2 (4.3) |
| - 65-69 | 40 (87.0) |
| - 70+ | 4 (8.7) |

The median time from RT completion to the latest available, post-RT CT was 43 months (IQR 32-57). Irradiated carotid shrinkage was observed in 78% and there was a statistically significant difference in the mean percent change (±SD) of the total irradiated versus spared carotid artery volumes, −7.0±9.0 vs. +3.5±7.2, respectively, (p<.0001).

The mean percentage changes in subvolumes of the carotid artery that was exposed to 50-55, 55-60, 60-65 and 75-70+ Gy in comparison to the mean percentages changes in the subvolumes of the contralateral carotid artery are presented in Table 2. No significant trend (dose-response relationship) was found on analysis of 5 Gy range subvolumes via one-way ANOVA, the differences in mean percent changes (±SD) for the 50-55, 55-60, 60-65, and 65-70+ Gy range subvolumes (irradiated minus spared) being −13.1±14.7, −9.8±14.9, −6.9±16.2, −11.7±11.1, respectively (p=0.217; Table 2 and Figure 2). Two patients (4%) had CVAs, both occurring in patients with greater % decrease in carotid artery volume in the irradiated versus the spared side. Furthermore, both patients who developed a CVA were white men treated with chemotherapy and a total of 66Gy of RT.

**Figure 2:**
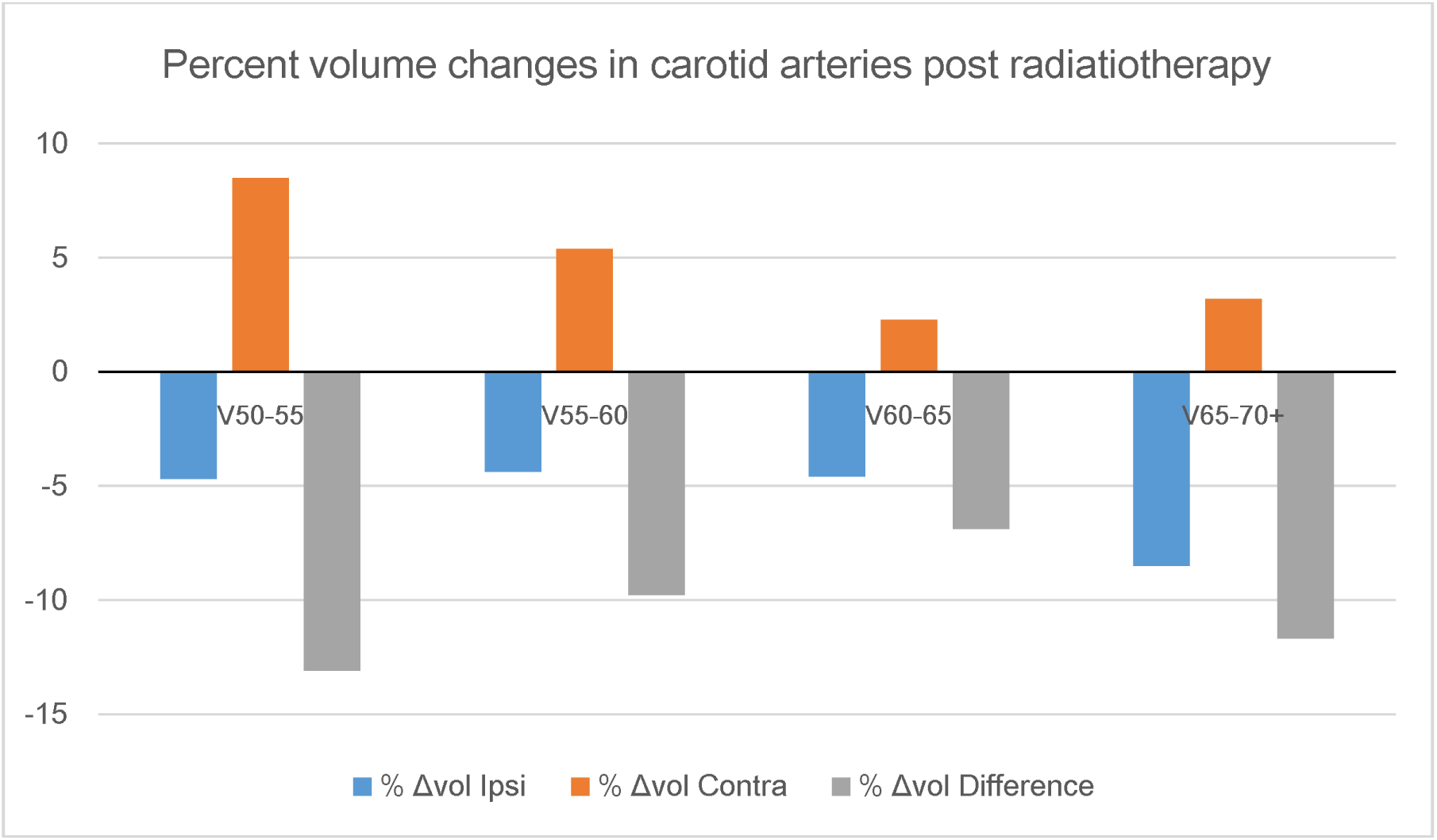
Mean percent volume change before and after radiation therapy in the ipsilateral (blue) and contralateral sides (orange) as well as mean difference in percent volume change (gray) in association with radiation dose volumetric parameters.

**Table 2.** Mean percent volume change before and after radiation therapy in the ipsilateral and contralateral sides as well as mean difference in percent volume change (ipsilateral minus contralateral) in association with radiation dose volumetric parameters.

| Subvolumes | Mean % Volume change (pre- and post-RT) in ipsilateral carotid artery (SD) | Mean % Volume change (pre- and post-RT) in contralateral carotid artery (SD) | Mean difference in % volume change (ipsilateral – contralateral) (SD) |
| --- | --- | --- | --- |
| V50-55 Gy | -4.7 (11.7) | 8.5 (17.5) | -13.1 (14.7) |
| V55-60 Gy | -4.4 (14.9) | 5.4 (16.5) | -9.8 (14.9) |
| V60-65 Gy | -4.6 (14.4) | 2.3 (12.8) | -6.9 (16.2) |
| V65-70+ Gy | -8.5 (10.6) | 3.2 (8.2) | -11.7 (11.1) |

No significant difference in overall carotid artery (ICA/CCA) volume between the treated and untreated sides were detected among patients who received systemic therapy versus those who did not (p>.05). All patients who received chemotherapy, were treated with 66 Gy of RT, while the mean dose of RT ± SD among patients treated with RT alone was 66.48 ±1.80 (p=0.34). Analysis by both receipt of systemic therapy and by 5 Gy range subvolumes via one-way ANOVA similarly did not reveal any statistically significant difference. There was not a statistically significant difference in the total carotid artery (ICA/CCA) volume change between current or former smokers versus nonsmokers, nor was there a significant difference when analyzing additionally by 5 Gy range subvolumes via one-way ANOVA. Finally, neck dissection was not associated with a statistically significant difference in overall carotid artery (ICA/CCA) volume via Wilcoxon test (p>.05), nor was there a significant difference via one-way ANOVA on analysis by 5 Gy range subvolumes (p>0.5).

## Discussion

The current study demonstrated that standard-of-care CT scans can be used to detect carotid artery volume changes following unilateral RT for early tonsillar SCC. We found that patients treated with greater than 50 Gy to the majority of the carotid artery had a significant reduction in total carotid artery volume (ICA/CCA). On examination for a dose-response relationship by 5 Gy ranges from 50-55, 55-60, 60-65, 65-70+ Gy, no significant differences were found, suggesting no further dose-response effect beyond 50 Gy. Although there may be a dose-response threshold under 50 Gy, we were not able to investigate that question in our dataset as very small, discontinuous volumes of the carotid received these lower doses.

CAS following RT is well-documented. Based on a systematic review of the literature, the prevalence of CAS was increased by 16-55% after RT; however the majority of studies were suboptimally matched, retrospective, and/or used historical radiation approaches.(11) Despite its prevalence, the pathogenesis of RICAD is still not completely understood.(12, 13) RICAD develops over years through typical atherosclerotic processes, but at a faster pace compared with unirradiated atherosclerotic arteries. (14) The interval from RT to symptomatic vascular changes ranges between several months to twenty years.(15, 16) For non-radiation related CAS, the most common sites are the carotid bifurcation and proximal segment of the internal carotid artery (ICA). This differs in RICAD, with the common carotid artery (CCA) being the most affected. (16, 17) RICAD sometimes extends beyond the radiation field, often to the proximal CCA and distal ICA, which are difficult to detect with US, but can be detected with CT. (16, 18)

Current routine methods for imaging the carotid arteries for determination of stenosis include duplex sonography, CT angiography (CTA), digital subtraction angiography, and magnetic resonance angiography.(19) Traditionally, measurement of the lumen has been the standard method of determining CAS on angiographic imaging studies, but also measurement of artery thickness, such as intima-media thickness (IMT) and the interadventitial diameter (IAD) have emerged as avenues for detection of subclinical atherosclerosis, as well as prognostic tools.(4, 20) IMT, which is determined by ultrasound, has been shown to be an early marker of radiation-induced carotid artery damage and has been utilized by the majority of studies examining RICAD. (12, 16, 21) IAD and whole vessel diameter enlargement, which can be measured on CTA, have been found to occur early in atherosclerosis, become exaggerated in the presence of vulnerable plaques, and were correlated to cardiac events and CVA.(20, 22–24) IMT and IAD only assess local changes and might not capture the whole extend of RICAD. Carotid artery volume estimation by CT is a 3-dimentional measure that includes the whole length of the extracranial carotid artery and can detect subsegmental changes especially in the setting of heterogeneous dose delivery patterns of RT.

Patients with OPC undergo evaluation with several CT scans of the head and neck area with contrast for cancer diagnosis and staging purposes, determination of therapy response or evaluation for cancer or cancer therapy related complications. (25) While the soft-tissue visualization optimized CT scans generally available for tonsillar cancer patients are not optimized for study of the carotid arteries, sufficient resolution is still present to accurately define the external diameter, as well as identify plaques. CT with contrast also allows for good resolution of the entire vascular tree from the CCA’s origin at the aorta to the ICA’s entry of the skull as compared to ultrasound.(4) In this study, we proved that standard-of-care oncologic surveillance soft-tissue visualization optimized CT scans with contrast can be used to successfully monitor whole carotid artery temporal volume changes in association with RT dose exposure. This approach enables the potential (re)use of clinically collected imaging studies to identify vulnerable patients at risk of CAS and its devastating associated morbidity and mortality. Additionally, in our cohort, three out of four patients had a decrease in their carotid artery volume, while an average increase in the volume of the spared carotid artery was noted. We hypothesize that this increase in the spared carotid artery volume might be compensatory to the decrease in the volume of the irradiated artery in order to maintain unchanged intracranial blood perfusion. This hypothesis needs to be further tests in future studies.

Two recent retrospective studies examined the relationship of radiation dose and risk of CAS (dose-response relationship) and reported increased risk with all dose-levels between 10-70 Gy.(8, 9) In our study, we did not find any significant difference in the percent volume change of the carotid artery in association with radiation dose for doses above 50 Gy divided in 5 Gy intervals. Lower doses could not be investigated as only small, discontinuous volumes of the carotids received doses <50 Gy. It is possible that there is a dose-response threshold under 50 Gy, as the current literature indicates, above which there is a significant increase in the risk of CAS, but with no further increase in the risk for dose groups above 50 Gy. If indeed the dose-response threshold is as low as 10Gy, that means that efforts to decrease RT dose for the treatment of OPC might not necessarily lead to a decrease in the incidence of RICAD, since RT dose in such low levels is ineffective.

Importantly, the lack of dose–response at therapeutic elective doses (i.e. >50Gy) suggests that for the foreseeable future, despite recent dose de-escalation applications (e.g. ECOG 3311 which prescribed minimum *50Gy* to radiated post-surgical cases),(26) the majority of head and neck RT patients are at risk for carotid artery injury at levels detected in the current study. Moreover, the epidemic of radiocurable HPV+ diseases, wherein the majority of patients can expect extended survival, often decades after therapy, suggest an unmet need for more careful surveillance of sub-clinical-to-chronic-to-symptomatic radiovascular toxicity. Consequently, additional efforts are needed to characterize and monitor whether the observed early changes in carotid lumen are indicative of later atherosclerotic disease states (e.g. asymptomatic carotid artery stenosis, plaque formation, TIA, CVA), and could serve as an early harbinger risk or as a risk stratification tool, easily integrated into standard CT surveillance imaging. Additionally, our data suggest that efforts to reduce carotid dose (e.g. unilateral radiotherapy,(27, 28), margin reduction,(29) or conformal techniques,(30, 31) are justifiable based on secondary imaging showing sub-acute vascular injury, and should be consistently considered when feasible if they do not compromise tumor control or oncologic recurrence risk.

In terms of the incidence rate of TIA/CVA, previous data has suggested that it measures approximately 5% in 5 years and 10% in 10 years following RT.(8) Our results demonstrated an incidence rate of 4% over a median follow up of 43 months, which is in line with the incidence rates reported in the literature. Both CVA s in our cohort occurred in patients that had irradiated carotid artery volume loss greater than on the spared side.

This study has several limitations, including the small sample size and retrospective nature with its inherent risk of bias. Furthermore, most patients in this study were men and white, which decreases generalizability of our findings. Despite the small sample size, we were still able to detect statistically significant changes in the main outcome reported in our study. Finally, our study does not include data on lipid levels and how lipid-lowering therapies such as statins would modify the change in carotid artery volumes.

## Conclusions

This study provides evidence that the volume of carotid artery significantly decreases following RT compared to the contralateral unirradiated carotid artery among patients with tonsillar cancer. Standard-of-care CT scans can be used to detect these changes and help risk stratify patients who will benefit from dedicated carotid artery imaging and frequent surveillance. Based on these findings, future studies could enable use of standard surveillance CT scans to screen for carotid artery stenosis in head and neck cancer survivors as well as lead to the creation of clinically meaningful mechanisms to modify the risk of carotid radiation vasculopathy and reduce radiation-associated morbidity and mortality.

## Data Availability

All data produced in the present study are available upon reasonable request to the authors.

## Acknowledgements

We would like to thank Aasheesh Kanwar from Willamette Valley Medical Center, McMinnville, OR for his valuable contribution to this study.

